# Impact of socio-environmental determinants and hygiene practices in tungiasis transmission in a vulnerable rural community

**DOI:** 10.1101/2025.06.05.25329050

**Authors:** Taiye Shade Olusegun-Joseph, Nur Juliani Shafie, Daniel Akinsanya Olusegun-Joseph, Muinah Adenike Fowora, Mohammed Akinlabi Rufai, Adedayo Michael Awoniyi, Monsuru A. Adeleke

**Affiliations:** Department of Science Laboratory Technology, Yaba College of Technology, Yaba, Lagos, Nigeria; Faculty of Science and Marine Environment, Universiti Malaysia Terengganu, Kuala Nerus, Terengganu, Malaysia; Department of Medicine, College of Medicine, Lagos University Teaching Hospital, Lagos, Nigeria; Molecular Biology and Biotechnology Department, Nigerian Institute of Medical Research, Yaba, Lagos, Nigeria; Department of Zoology, Osun State University, Osogbo, Osun State, Nigeria; Institute of Collective Health, Federal University of Bahia, Salvador - BA, Brazil

**Author notes:** **Correspondence to:** TS Olusegun-Joseph - /NJ Shafie. These authors contributed equally (Adedayo Michael Awoniyi, Monsuru A. Adeleke).

**Keywords:** *Tunga* spp., Ectoparasite, Jigger, Prevention of tungiasis, Neglected Tropical Disease

## Abstract

**Background:** Tungiasis is a neglected tropical disease (NTD) caused by the penetration of adult female *Tunga* fleas into the skin of humans and animals. The disease disproportionately affects individuals in tropical and subtropical regions, particularly in sub-Saharan Africa, South America and Caribbean, where poverty, inadequate sanitation and limited healthcare services facilitate transmission, especially among vulnerable populations.

**Objective:** To evaluate the association between socio-environmental factors and personal hygiene practices with tungiasis prevalence among residents of rural communities in the periphery of Badagry, Lagos State, Nigeria, across the rainy and dry seasons from September 2021 – April 2022.

**Methods:** A cross-sectional survey was conducted, targeting at least 423 participants per sampling season. Following ethical approval and informed consent, semi-structured questionnaires were administered to collect demographic, socioeconomic, environmental and hygiene-related data. Physical examinations, performed by a qualified medical practitioner, identified tungiasis cases based on previously validated criteria. Descriptive statistics were used to summarize the data, while the Chi-square test assessed associations between infection status and categorical variables. A multivariate generalized linear model was used to identify key predictors of tungiasis infection.

**Results:** Overall tungiasis prevalence of 5.3% (54/1020) among study participants. While infection rates were slightly higher in males (27/410; 6.6%), the difference was not statistically significant. Most infected participants (96.3%) had lower education levels and exhibited unsatisfactory hygiene practices. The Chi-square test identified residing in a household with an earthen floor as the strongest variable correlated with tungiasis [χ^2^= 63.2, *p* = <**0.001**]. However, the final model, revealed seasonality -rainy season (OR: 4.80 [2.33–10.82], *p = **<*****0.001**), frequent rat sightings (OR: 3.04 [1.40–7.29], *p =* **0.008**) and sleeping on a mattress placed directly on the floor (OR: 2.60 [1.18–6.28], *p =* **0.024**) as the most significant determinants of tungiasis infection.

**Conclusions:** Our findings highlight the complex interplay between socioeconomic status, environmental characteristics, hygiene practices and seasonal variations in shaping tungiasis prevalence within a vulnerable rural community. Key predictors of infection, that is, seasonality, frequent rat sightings and poor sleeping conditions, underscore the need for targeted interventions to prioritize improving personal and household hygiene, implementing sustainable rodent control measures and raising community awareness about tungiasis prevention. These measures will not only help reduce tungiasis prevalence but also mitigate the burden of other zoonotic diseases that thrive under similar socio-environmental conditions.

## Introduction

Tungiasis is a parasitic skin disease caused by adult female sand fleas, primarily *Tunga penetrans*, and less frequently *T. trimamillata* [1]. The fleas burrow into the skin of humans and animals, commonly affecting the hands, knees, heels and soles [2, 3] The disease is known by various local names, including jiggers, chigoe, pique and nigua [2, 3]. Tungiasis progresses through five stages: 1) penetration-where the flea partially embeds itself in the skin, causing itching as the primary symptom; 2) early hypertrophy-here, a central brown spot (0.5-2 mm) appears, surrounded by an erythematous areas; 3) maximal hypertrophy-the lesion enlarges (3-10 mm in diameter) with a characteristic white circular zone and a small central black dot. Symptoms include erythema, edema, tenderness, heat, pain, intense itching and flaking of the corneal layer around the lesion; 4) initial involution-here, the lesion begins to shrink, with the hypertrophic zone reducing approximately three weeks after penetration; and 5) expulsion-where the parasite is expelled from the body, falling onto the soil or floor [4, 5].

The spread of parasite eggs in the environment primarily occurs through domestic animals, which serve as key reservoirs. These include dogs, cats, pigs, cattle and rodents [4, 6]. The severity of the disease is directly correlated with the number of embedded fleas, with heavily infested individuals facing an increased risk of developing open wounds and secondary infections. Beyond the localized skin damage, tungiasis can lead to severe systemic complications, including anemia and tetanus, both of which if untreated could result in fatal outcomes [7, 8]. Also, medical complications such as inflammation, ulcerations, fibrosis, lymphangitis, gangrene, and sepsis may also arise due to secondary infections following flea infestation [9]. This neglected tropical disease (NTD) is widespread in tropical and subtropical regions, affecting millions of people across South America, the Caribbean and sub-Saharan Africa [10, 11, 12].

Studies on tungiasis risk factors have consistently highlighted that factors such as overpopulation, illiteracy, inadequate public healthcare facilities, traditional beliefs, poor health-seeking behavior and sanitation deficiencies associated with poverty all contribute to its high prevalence [13, 14, 15]. Additionally, poor hygiene and environmental conditions significantly increase the risk of infection. For example, poor household sanitation, living in dwellings with earthen floors, improper animal care and poor personal hygiene such as wearing dirty clothes and walking barefoot, may further elevate susceptibility to tungiasis [2, 6].

The coastal community used for this study is known as a tungiasis-prone community, probably due to its precarious socio-environmental conditions and inadequate healthcare facilities [16, 17]. However, there remains a significant gap in comprehensively evaluating the interplay between socioeconomic factors, environmental characteristics and hygiene practices in driving tungiasis prevalence, particularly in vulnerable rural environments. Globally, studies evaluating these interconnections remain scarce [18] and even more limited in Nigeria and other low- and middle-income countries (LMICs). Addressing this knowledge gap is essential, as a deeper understanding of these determinants is crucial for designing evidence-based and region-specific interventions to mitigate tungiasis burden, especially in high-risk and vulnerable communities.

This study aims to evaluate the association between socio-environmental and personal hygiene practices with tungiasis prevalence among residents of a vulnerable rural community in the periphery of Badagry, Lagos State, Nigeria. The results of this study should provide policymakers with valuable insights for designing sustainable interventions to reduce tungiasis and other related zoonotic diseases transmission in rural communities, particularly those located in LMICs.

## Methods

### Study Area

This study was conducted in remote rural communities in the periphery of Badagry (6.41622°N; 2.89077°E), Badagry Local Government Area (LGA) of Lagos State, Nigeria. Badagry is a coastal region with an estimated population of approximately 327,400 based on the 2016 projection [19]. The region has been reported as a tungiasis-prone area [14, 17, 20, 21].

The climate in Badagry is tropical, with an average daily temperature of approximately 30°C and annual mean rainfall of about 1532 mm. The region experiences two distinct seasons: the rainy season, which spans from April to October and the dry season, occurring between November and March [21]. The primary occupations of residents include fishing, farming, trading and to a lesser extent, civil service. The region is predominantly composed of Yoruba and Egun ethnic groups, with Christianity being the dominant religion. Socioeconomic and infrastructural challenges characterize the region, including inadequate access to potable water, erratic electricity supply and unsatisfactory healthcare facilities. Additionally, poor environmental conditions are prevalent, including substandard housing (Figure 1A), open sewage systems, improper waste disposal and precarious garbage collection services. Frequent household rodent infestation and the free roaming of domestic animals around residences further contribute to the unsanitary conditions of the region [21].

**Figure 1:** (A) Illustration of a typical hut of some participants; (B) Common local treatment of infected individuals, through the application of oil on the inflamed foot; (C) A severely deformed heel of an infected participant, showing multiple inflamed tungiasis lesions; and (D) A traditional method of extracting embedded fleas from the foot of an infected participant using thorn. The figure photographed by Taiye Shade Olusegun-Joseph, Department of Science Laboratory Technology, Yaba College of Technology, Yaba, Lagos, Nigeria

### Study Design

#### Sample size considerations and eventual study population

The sample size was estimated using a slightly modified Cochran’s formula for estimating a population proportion [22]. Briefly, the formula is as follows:

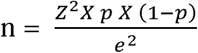

Where,

n = required sample size to estimate the population proportion with a desired level of precision; Z = z-score corresponding to the desired confidence level (1.96 for a 95% confidence interval);

P = estimated proportion of the population possessing the characteristic of interest; and e = acceptable margin of error (degree of accuracy).

Here, we used a Z-score of 1.96 for a 95% CI, an estimated proportion (p) of 45.2% based on Ugbomoiko et al., [14] and a margin of error (e) of 5% to calculate the initial sample size (n), yielding sample size of 380.6 approximately 381 participants.

Adjusting the sample size (n_f_) for 10% non-response:

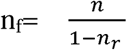

Where n_f_ = adjusted sample size and n_r_ = non-response rate (10%)

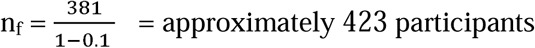

As a result, we targeted a minimum of 423 participants for each sampling season. Specifically, consenting heads of households were invited to participate in the study by responding to a semi-structured questionnaire.

#### Eligibility criteria

The inclusion and exclusion criteria were determined based on the duration of residence within the community. Individuals who had lived in the neighborhood for at least three months before the study commenced and who spent a minimum of four nights per week in the community were eligible to participate in the study. Exclusion criteria included visitors and individuals who had resided in the community for less than three months before the commencement of the study. Additionally, only households and individuals who provided informed consent (written/verbal) were included in the study.

#### Data collection and procedures

The survey was conducted from 27^th^ September 2021 – 30^th^ April 2022, covering both the rainy and dry seasons. Socioeconomic, environmental and hygiene-related variables were collected through semi-structured questionnaires, which gathered data on demographics and socioeconomic status, environmental conditions, domestic animal ownership, rat sightings and hygiene behaviors.

To determine infected and non-infected individuals, a qualified medical practitioner conducted thorough physical examinations to identify signs of tungiasis. The diagnosis was based on the presence of clustered nodules, white patch with a black dot [4] and or embedded *Tunga* flea (flea lesions - Figure 1B&C) on the legs, feet, hands and arms of participants, following validated protocols by Girma et al., [23] and Jorga et al., [24]. For consenting individuals, embedded fleas were extracted using a sterile hypodermic needle under sterile conditions by the experienced medical practitioner as a safe alternative to the traditional method of using thorns for flea removal (Figure 1D).

### Statistical data analysis

We pseudo-anonymized all individual data to ensure participants’ privacy upon data cleaning. We performed descriptive analysis to estimate the frequency and percentage of total, infected and non-infected participants. We used the Chi-square test of independence to evaluate the differences between the mean of tungiasis infection status and categorical variables, including demographic and socioeconomic characteristics, environmental factors, presence of domestic animals and household rat infestation and resident hygiene behaviors. Statistical significance was set at *p <* 0.005.

Before constructing the multivariate generalized linear models (GLM), we performed a univariate analysis to identify potential predictors of tungiasis infection and reduce the number of variables included in the final model. The univariate analysis evaluated the relationships between tungiasis infection status (infected versus non-infected) and a range of explanatory variables, including: sampling season (dry vs rainy); age (continuous variable); gender; religion; education level; feet hygiene (assessed through inspection to determine individual neatness); occupation; clothes hygiene (evaluated by direct inspection and self-reported washing frequency of participants clothes); shoe-wearing habits; presence of domestic animals in the household or compound; sleeping place; school/work attendance frequency per week; household rat infestation (evaluated based on participant reported sightings); residence with unpaved floor (earthen floor); households with unplastered walls and number of household residents.

Variables with *p*-values ≤ 0.2 in the univariate analysis, along with those previously identified as relevant to tungiasis transmission were included in a preliminary multivariate model. We used this technique to prevent the premature exclusion of potentially important predictors, as adopting the conventional *p* < 0.05 threshold at this stage could lead to the omission of key predictors [25]. To assess multicollinearity, we calculated the variance inflation factor (VIF) and excluded variables with VIF > 3 in the analysis [26]. The final model was determined using a mixed forward and backward stepwise model selection approach using Akaike’s Information Criteria (AIC). The simplest model with ΔAIC < 2 was selected following the parsimony principle [27]. All statistical analyses were performed using R software, version 4.4.1 [28].

### Ethical considerations

We ensure that all methods were conducted following relevant guidelines and regulations and reported following the ARRIVE guidelines. Ethical approval for this study was obtained from the Nigeria Institute of Medical Research Ethics Committee (approval number IRB/21/053). Additionally, we secured authorization from the Badagry Local Government Area Public Health Board and obtained permission from the community leader of the study community. Informed oral/written consent was obtained from household heads after thoroughly explaining the study’s purpose and only consenting participants were enrolled.

## Results

### Sociodemographic and environmental characteristics

Table 1 summarizes the socio-environmental characteristics of the study participants. A total of 1020 participants were interviewed and examined for tungiasis, with response rates ranging from 997 to 1020 (97.7% −100 %). Data collection was homogeneously distributed across seasons within the study community. The mean age of participants was 38 ± 15.5 years, and households had an average of 5 ± 4.5 residents. Females comprised the majority 610 (59.2%) of participants.

**Table 1:** Demographic profile, socio-environmental characteristics and hygiene behaviors of residents in a vulnerable rural community on the periphery of Badagry, Lagos State, Nigeria, participating in a tungiasis assessment study.

| Variables | All, n (%) | Tungiasis infection |  | Statistical test |
| --- | --- | --- | --- | --- |
|  |  | Yes, n (%) | No, n (%) |  |
| **Season** | | | | $\chi^2=23.9, p < 0.001$ |
| Dry | 528 (51.8) | 10 (1.9) | 518 (98.1) |  |
| Rainy | 492 (48.2) | 44 (8.9) | 448 (91.1) |  |
| **Gender** | | | | $\chi^2=1.89, p = 0.172$ |
| Female | 610 (59.8) | 27 (4.4) | 583 (95.6) |  |
| Male | 410 (40.2) | 27 (6.6) | 383 (93.4) |  |
| **Education level** | | | | $\chi^2=5.85, p = 0.210$ |
| Uneducated | 240 (23.5) | 11 (4.6) | 229 (95.4) |  |
| Primary | 343 (33.6) | 26 (7.6) | 317 (92.4) |  |
| Secondary | 389 (38.1) | 15 (3.9) | 374 (96.1) |  |
| University | 32 (3.1) | 1 (3.1) | 31 (96.7) |  |
| Others | 13 (1.3) | 1 (7.7) | 12 (92.3) |  |
| Missing | 3 (0.3) | 0 (0) | 3 (100) |  |
| **Main occupation** | | | | $\chi^2=26.3, p <0.001$ |
| Farmer | 53 (5.2) | 2 (3.8) | 51 (92.2) |  |
| Fishermen | 154 (15.1) | 8 (5.2) | 146 (94.8) |  |
| Salary earner | 9 (0.9) | 0 (0) | 9 (100) |  |
| Trader | 411 (40.3) | 20 (4.9) | 391 (95.1) |  |
| Student | 94 (9.2) | 15 (16) | 79 (84) |  |
| Casual worker | 2 (0.2) | 0 (0) | 2 (100) |  |
| Artisan | 48 (4.7) | 1 (2.1) | 47 (97.9) |  |
| Others | 240 (23.5) | 7 (2.9) | 233 (97.1) |  |
| Missing | 9 (0.9) | 1 (11.1) | 8 (88.9) |  |
| **Feet hygiene** | | | | $\chi^2=39.9, p <0.001$ |
| Clean | 256 (25.1) | 5 (2) | 251 (98) |  |
| Fairly clean | 620 (60.8) | 26 (4.2) | 594 (95.8) |  |
| Dirty | 144 (14.1) | 23 (16) | 121 (84) |  |
| **Clothes hygiene** | | | | $\chi^2=30.5, p <0.001$ |
| Clean | 348 (34.1) | 8 (2.3) | 340 (97.7) |  |
| Fairly clean | 584 (57.3) | 31 (5.3) | 553 (94.7) |  |
| Dirty/unwashed | 88 (8.6) | 15 (17) | 73 (83) |  |
| **Shoe wearing** | | | | $\chi^2=1.65, p <0.437$ |
| Always | 197 (19.3) | 8 (4.1) | 189 (95.9) |  |
| Irregular | 501 (49.1) | 25 (5) | 476 (95) |  |
| Never | 322 (31.6) | 21 (6.5) | 301 (93.5) |  |
| **Sleeping place** | | | | $\chi^2=32.8, p <0.001$ |
| Mattress on bed | 528 (51.8) | 30 (5.7) | 498 (94.3) |  |
| Mattress on floor | 369 (36.2) | 10 (2.7) | 359 (97.3) |  |
| Mat on floor | 109 (10.7) | 9 (8.3) | 100 (91.7) |  |
| Others | 14 (1.4) | 5 (35.7) | 9 (64.3) |  |
| **Possess animal** | | | | $\chi^2=0.01, p = 1.000$ |
| No | 376 (36.9) | 20 (5.3) | 356 (94.7) |  |
| Yes | 644 (63.1) | 34 (5.3) | 610 (94.7) |  |
| **Rat sightings** | | | | $\chi^2=4.79, p = \mathbf{0.029}$ |
| No | 359 (35.2) | 11 (3.1) | 348 (96.9) |  |
| Yes | 661 (64.8) | 43 (6.5) | 618 (93.5) |  |
| **Earthen floor** | | | | $\chi^2=63.2 p < \mathbf{0.001}$ |
| No | 847 (83) | 23 (2.7) | 824 (97.3) |  |
| Yes | 173 (17) | 31 (17.9) | 142 (82.1) |  |

Note: *n* represents the number of variables, with the corresponding percentage in parentheses

Regarding education levels, 389 (38.1%) had secondary/high school education, 343 (33.6%) had primary/basic school education and 240 (23.5%) had never received formal education. In terms of occupation, 411 (40.3%) were traders, followed by 154 (15.1%) fishermen. Additionally, 173 (17%) participants resided in households with earthen flooring.

### Hygiene practices and house conditions

The study participants’ individual hygiene and housing conditions are presented in Table 1. Among the 1020 participants, only a minority, 256 (25.1%) had clean feet, while the remaining participants exhibited slightly or visibly dirty feet. Similarly, only 348 (34.1%) maintained satisfactory clothing hygiene, characterized by regularly washing and wearing satisfactorily clean clothes. The habits of shoe wearing varied among participants, for example, the majority 501 (49.1%) reported irregular shoe use, while 322 (31.6%) stated they had never worn shoes, habitually walking barefoot. Nearly half of the participants 478 (46.9%) sleep on a mattress or mat (a locally made alternative crafted from plant or rubber materials) placed directly on the floor. Household environmental conditions revealed 661 (64.8%) of participants frequently sighting rats in and around their households, indicating high levels of household rat infestation. Also, 644 (63.1%) reported rearing various domestic animals, including chickens, goats, sheep, cats and goats within their compounds.

### Prevalence and determinants of tungiasis

As presented in Table 1 above, tungiasis was detected in 54 out of the 1020 participants, representing an overall prevalence of 5.3% across the study community. The prevalence was slightly non-significantly higher among male participants, with 27 out of 410 males (6.6%) testing positive, accounting for 50% of all cases. Also, majority of affected participants (52 out of 54; 96.3%) had primary, secondary or no formal education.

To identify factors potentially influencing tungiasis prevalence among study participants, using tungiasis infection status (infected and non-infected) in a vulnerable rural community, thirteen variables with *p*-values ≤ 0.20 or those previously identified as relevant to tungiasis transmission from the initial univariate analysis were included in the potential final model (supplementary Table I). Of these variables, six key variables (namely: season; age; feet hygiene; sleeping place; rat sightings and earthen floor) were retained in the final model (Figure 2) with an Akaike Information Criterion (AIC) of 289.9, representing the model with the fewest variables.

**Figure 2:** Results of the generalized linear model of variables associated with tungiasis among study participants in a vulnerable rural community in the periphery of Badagry, Lagos State, Nigeria.

The Chi-square test suggests residing in a household with an earthen floor as the strongest variable correlated with tungiasis [χ2=63.2 *p* = <**0.001**]. Other significant factors included participants with unclean feet [χ^2^=39.9, *p* = <**0.001**], unsatisfactory sleeping conditions [χ^2^=32.8, *p* = <0.001], unclean clothes [χ^2^=30.5, *p* = <**0.001**], being a trader [χ^2^=26.3, *p* = <**0.001**] and the sampling season [χ^2^=23.9, *p* = <**0.001**]. Similarly, participants who owned domestic animals and those reporting frequent rat sightings reveal a strong correlation with tungiasis infection.

However, results from the final model (Figure 2) showed that residing in a household with an earthen floor (OR: 0.10 [0.04–0.19], *p = **<*****0.001**) and having dirty feet (OR: 0.15 [0.04–0.44], *p =* **0.001**) were not the strongest predictors of exposure. In contrast, participants who: sleep on a mattress placed on the floor (OR: 2.60 [1.18–6.28], *p =* **0.024**), reported frequent rat sightings (OR: 3.04 [1.40–7.29], *p =* **0.008**) and surveyed during rainy season (OR: 4.80 [2.33–10.82], *p = **<*****0.001**) had significantly higher odds of tungiasis infection. Notably, the rainy season and frequent rat sightings emerged as the most significant predictors, with individuals surveyed during the rainy season being 4.8 times more likely to have tungiasis and those reporting frequent rat sightings being 3 times more likely to be infected compared to their counterparts surveyed in the dry season and those without frequent rat sightings.

## Discussion

Studies evaluating the association between socio-environmental factors, personal hygiene behaviors and tungiasis prevalence in vulnerable rural communities, particularly in LMICs, remain scarce. Here, our results highlight the complex interplay between socioeconomic status, environmental characteristics, hygiene practices and seasonal variations in influencing tungiasis prevalence within the study community. Notably, we observed that seasonality and frequent rat sightings emerged as the most significant predictors of tungiasis infection, underscoring the role of environmental factors in disease transmission. Although synanthropic animals like *Rattus* species and domestic animals such as pigs, dogs and cats are well documented hosts of the vectors and fleas [21, 29, 30], the higher prevalence observed during the rainy season may be attributed to the prolonged wet conditions in the coastal study area, which somehow aligns with the findings by Harvey et al. [31], who reported no significant seasonal variation in tungiasis prevalence due to the persistent all year rainfall in their study community. However, our findings differ from some previous studies where prevalence tended to peak slightly during the dry season [32, 33]. These disparities suggest that the seasonal dynamics of tungiasis may vary depending on local climate patterns, variations in the current climatic conditions and ecological factors, emphasizing the need for region-specific disease control strategies.

The mean age of infected participants (29 ± 15 years) was lower than that of non-infected participants (39 ± 15 years), suggesting that younger participants may be at higher risk of tungiasis. This increased vulnerability may stem from their higher engagement in environmentally related activities, leading to increased exposure to key predictors [2, 3]. Although the majority of study participants were female (610, 59.2%), tungiasis prevalence was slightly higher (but non-significant) among males. This disparity may be attributed to higher participation in outdoor activities that elevate their exposure risks, mirroring the trend observed among younger participants [3]. The low education levels of many infected participants likely reflect their poor socioeconomic status, which may limit their ability to reside in better-developed communities with improved sanitation services, healthcare access and disease prevention programs [10, 18, 34,35]. While this study did not specifically assess knowledge of tungiasis transmission and prevention strategies, participants’ limited educational background suggests a possible lack of awareness regarding the disease cycle and effective prevention measures. This is further evidenced by participants’ reported traditional treatment practices, where the majority indicated using thorns to extract embedded fleas rather than seeking appropriate medical care (Figure 1D). The socioeconomic challenges of infected individuals are also reflected in their occupational profiles, as many are engaged in fishing, farming and trading activities that could increase exposure to sand flea-infested environments. Similarly, their poor living conditions (Figure 1A), such as earthen floors and sleeping on mats or mattresses placed directly on the floor, may further heightened their vulnerability. Their hygiene practices, including walking barefoot and wearing dirty clothes, may also increase their exposure to sand fleas and consequently tungiasis infection [10, 36, 37, 38], highlighting the need for targeted health education, improved sanitation and community-based interventions to mitigate the disease risk, particularly in vulnerable populations.

The personal hygiene behaviors and housing conditions of study participants highlight important risk factors that may contribute to tungiasis transmission. Poor foot hygiene was common among participants, with only a small minority maintaining clean feet, while the majority also frequently walked barefoot, exposing participants to infected surfaces and sand fleas [10, 37]. Poor sleeping conditions were also common, as many participants placed their mattresses or mats directly on the floor, further increasing contact with flea-infested surfaces. Similarly, poor clothing hygiene was widespread, potentially reflecting a broader lack of access to potable water, sanitation and hygiene resources. The lack of these essential services not only compromises personal hygiene but also contributes to increased vulnerability to zoonotic diseases, including tungiasis [36]. The housing conditions within the community created a favorable environment for tungiasis transmission. The majority of study participants reported owning domestic animals, while a high proportion experienced frequent household rat infestation, which may increase exposure risk given these animals may serve as the primary vectors of *Tunga* species with previous studies also establishing that households with high densities of domestic animals and rodent infestations tend to harbor increased flea populations, as a result elevating tungiasis transmission risks [21, 36, 39]. These findings underscore the need for integrated disease control strategies to mitigate tungiasis risks and improve overall health conditions of the vulnerable populations.

The findings here revealed an overall tungiasis prevalence of 5.3% among the study participants, indicating an ongoing burden of the disease within the community. Although, this prevalence appears moderate compared to reports from other endemic regions, where higher prevalence such as 34% in a vulnerable community in Brazil [34], 52% among children in a rural district of Ethiopia [24] and 62% among school-aged children in a rural county of Kenya [35]. The prevalence observed here is higher than the 1.1% reported in a similar coastal area of Kenya [40] and falls within the 1.6% to 54.8% range reported in a systematic review of tungiasis prevalence in Brazil [41] highlighting the persistent burden of the disease in the study area.

Although the Chi-square analysis initially suggested that residing in a household with an earthen floor was the strongest predictor of tungiasis infection, our multivariate modeling refined this correlation, indicating other predictors as more significant predictors of infection risk. In the final model, seasonality, poor sleeping conditions and frequent rat sightings emerged as the most significant determinants of tungiasis infection. Participants reporting frequent rat sightings may reflect broad unsatisfactory household hygiene [42, 43], increasing their susceptibility to zoonotic diseases such as tungiasis through greater exposure to the parasite’s host and flea-infested surfaces [10, 36, 37]. Given that both analyses suggest poor sleeping conditions and frequent rat sightings are significantly correlated with tungiasis infection, future interventions targeted at reducing tungiasis should consider advocating improved sleeping conditions of participants and sustainable pest rodent management measures to enhance the general participants’ well-being. Similarly, the strong correlation between seasonality and infection prevalence underscores the need for targeted control during peak transmission, as increased moisture might create favorable conditions for the flea parasite’s survival and reproduction in certain regions, influencing higher infestation among residents [44, 45].

While our study explored a broad range of socio-environmental and behavioral factors potentially correlated with tungiasis infection, allowing for a detailed understanding of the disease transmission dynamics, we acknowledge certain limitations. A longitudinal study would have further strengthened our findings by allowing a more robust assessment of causality between the identified predictors and tungiasis infection. Also, while our results suggest that household rat infestations and domestic animal ownership may contribute to infection risk, we were unable to directly assess flea burden in the environment or on hosts through entomological surveys. Such an approach could have provided stronger empirical evidence of transmission pathways. Future research incorporating environmental flea assessments would offer valuable insights into the role of synanthropic and domestic animals in sustaining tungiasis transmission. Despite these limitations, our study has several notable strengths. The use of both univariate and multivariate analyses strengthens the robustness and reliability of our findings by identifying the most significant predictors of infection. Also, the relatively large and diverse sample size increases the statistical power of our analyses, improving the generalizability of our findings to other similar vulnerable rural communities. Importantly, by evaluating tungiasis prevalence across different seasons, our study provides critical region-specific insights into seasonal variations in transmission, which is essential for designing region-specific targeted intervention strategies to mitigate infection risk during peak transmission periods.

## Conclusion

In conclusion, our results show the complex interplay between socioeconomic status, environmental characteristics, hygiene behaviors and seasonal factors in shaping tungiasis prevalence within a vulnerable rural community. Specifically, poor sleeping conditions, frequent rat sightings and seasonality emerged as the principal predictors of infection risk. These suggest that effective interventions should consider promoting personal and household hygiene, sustainable pest rodent control measures and increasing community awareness of tungiasis transmission pathway and prevention strategies. Beyond reducing tungiasis prevalence, these interventions could also mitigate the burden of other zoonotic infections that thrive in similar environmental and socioeconomic settings, especially during peak transmission periods. To ensure sustainability and local acceptance, efforts to address the identified predictors here should prioritize community-led initiatives, leveraging the active involvement of research groups, public health authorities and other key stakeholders. By fostering collaborative, culturally appropriate interventions, we can encourage disease prevention and enhance the general well-being of at-risk populations, especially in endemic regions.

## Data Availability

All data produced in the present study are available upon reasonable request to the authors

## Acknowledgement

Open access funding provided by Universiti Malaysia Terengganu, Kuala Nerus, Terengganu, Malaysia. The authors appreciate community leaders and the residents for their cooperation during the study.

## Author contribution

TS Olusegun-Joseph, NJ Shafie, AM Awoniyi & MA Adeleke conceptualized the project; TS Olusegun-Joseph & DA Olusegun-Joseph collected the data; TS Olusegun-Joseph, & AM Awoniyi analyzed the data; TS Olusegun-Joseph, DA Olusegun-Joseph, MA Fowora, MA Rufai, AM Awoniyi & MA Adeleke developed the methodology; MA Fowora, MA Rufai & MA Adeleke supervised the project; TS Olusegun-Joseph, NJ Shafie & AM Awoniyi wrote the original draft of the manuscript; and TS Olusegun-Joseph, NJ Shafie, AM Awoniyi & MA Adeleke reviewed and edited the manuscript.

